# Evaluation of Patient-Level Retrieval from Electronic Health Record Data for a Cohort Discovery Task

**DOI:** 10.1101/19005280

**Authors:** Steven D. Bedrick, Aaron M. Cohen, Yanshan Wang, Andrew Wen, Sijia Liu, Hongfang Liu, William R. Hersh

## Abstract

**Objective:** Growing numbers of academic medical centers offer patient cohort discovery tools to their researchers, yet the performance of systems for this use case is not well-understood. The objective of this research was to assess patient-level information retrieval (IR) methods using electronic health records (EHR) for different types of cohort definition retrieval.

**Materials and Methods:** We developed a test collection consisting of about 100,000 patient records and 56 test topics that characterized patient cohort requests for various clinical studies. Automated IR tasks using word-based approaches were performed, varying four different parameters for a total of 48 permutations, with performance measured using B-Pref. We subsequently created structured Boolean queries for the 56 topics for performance comparisons. In addition, we performed a more detailed analysis of 10 topics.

**Results:** The best-performing word-based automated query parameter settings achieved a mean B-Pref of 0.167 across all 56 topics. The way a topic was structured (topic representation) had the largest impact on performance. Performance not only varied widely across topics, but there was also a large variance in sensitivity to parameter settings across the topics. Structured queries generally performed better than automated queries on measures of recall and precision, but were still not able to recall all relevant patients found by the automated queries.

**Conclusion:** While word-based automated methods of cohort retrieval offer an attractive solution to the labor-intensive nature of this task currently used at many medical centers, we generally found suboptimal performance in those approaches, with better performance obtained from structured Boolean queries. Insights gained in this preliminary analysis will help guide future work to develop new methods for patient-level cohort discovery with EHR data.

## INTRODUCTION

Many academic medical centers offer patient cohort discovery to their researchers to facilitate clinical research, usually including electronic health record (EHR) data [1, 2]. A number of systems are available to facilitate this task, such as i2b2 [3, 4] and TriNetX [5]. However, the performance of systems and algorithms for this EHR use case is not well-studied. It has been shown that typical review of patients for study eligibility is a labor-intensive task, and that automated preprocessing of lists of patients may reduce human time and effort for selection of cohorts [6-8].

One challenge for evaluating this use case is the lack of test collections that include data, clinical study descriptions, and relevance judgments for retrieved patients, a problem that has hindered many types of research using EHR data, even in the modern era of ubiquitous EHR adoption [9]. A major barrier has been the challenge of protecting privacy of the patients from whom the records are from and institutional hesitancy to making such data widely available for informatics research, even in de-identified form [10]. This is especially so for use cases involving processing of textual data within records, especially those used on the scale of information retrieval experiments where corpora of thousands to millions of patient records are typically desired.

There are two EHR record collections that have been publicly available, one from the University of Pittsburgh Medical Center (UPMC) [11] and the other the Medical Information Mart for Intensive Care-III (MIMIC-III) from the Massachusetts Institute of Technology [12]. Among the uses of the UPMC corpus has been a cohort retrieval for clinical research studies task in a challenge evaluation as part of the annual Text Retrieval Conference (TREC). The TREC Medical Records Track ran in 2011 and 2012, attracting 29 and 24 academic and industry research groups respectively [13, 14]. Using the University of Pittsburgh collection containing 17,264 encounters containing 93,551 documents (some of which included ICD-9 diagnosis codes, laboratory results, and other structured data), a total of 34 and 47 topics respectively by year were developed and relevance judgments performed based on pooled results from participating research groups using the “Cranfield paradigm” common to information retrieval (IR) evaluation research [15]. The judgments were performed by physicians enrolled in biomedical informatics educational programs.

Methods found to lead to improved retrieval performance included several domain-specific enhancements on top of word-based queries, including vocabulary normalization specific to the clinical domain, synonym-based query expansion from medical controlled terminology systems such as the Unified Medical Language System (UMLS) Metathesaurus, and recognition of negation [16]. Follow-on research with the test collection found continued improvement in performance from approaches such as query expansion for additional clinical and other corpora [17] as well as use of learning-to-rank methods [18].

One limitation of the TREC Medical Records Track was a limitation of the UPMC corpus, which was retrieval at the encounter (e.g., hospital or emergency department visit) and not the patient level. This was due to the de-identification process that broke the links across encounters, a process that also obscured various protected health elements, such as dates, geographic locations, and provider identifiers. Encounter-level retrieval data sets prohibit applying expert judgement and therefore evaluation at the patient level, which is the goal of cohort retrieval. Nonetheless, the TREC Medical Records Track did provide a data set for information retrieval and biomedical informatics researchers to compare different approaches to identifying patient cohorts for recruitment into clinical studies. Unfortunately, the UPMC corpus has been withdrawn from public use (Wendy Chapman, personal communication).

Outside of the TREC Medical Records Track, few other evaluations of cohort retrieval have been carried out and published. Some are limited by being document- or encounter-based, or focus on broadly defined cohorts that may be too general for the clinical research recruitment use case. One analysis using the MIMIC-III corpus looked at two straightforward clinical situations and found accurate retrieval with both structured data extraction and the use of natural language processing (NLP) [19]. Another recent approach employed word embeddings and query expansion to define patient cohorts, although used only structured EHR data [20]. The 2018 National NLP Clinical Challenges (n2c2) had a shared task devoted to cohort selection for clinical trials but focused on the complementary task of finding inclusion criteria of clinical trials as opposed to patient-level retrieval [21].

Another thread of work has focused on making querying easier to carry out, typically through development of natural language or other structured interfaces to the patient data [22-25]. Other approaches focus on normalizing semantic representation of patient data within the EHR itself and applying deep learning to non-topical characteristics of studies and researchers [27]. A related area to cohort discovery is patient phenotyping, one of the goals of which is to identify patients for clinical studies [28-30]. However, the cohort discovery use case has some differences, as some studies have criteria beyond phenotypic attributes, such as age, past treatments, diagnostic criteria, and temporal considerations.

In 2014, Oregon Health & Science University (OHSU) and Mayo Clinic launched a project to use raw (i.e., not de-identified) EHR data to perform research in parallel (i.e., able to share methods and systems but not data). The OHSU data set has been previously described [31], and this paper reports the first results using this data set along with evaluation at the patient level. The Mayo Clinic has reported some of its work, although its retrieval output and relevance judgments were at the encounter level and not the patient level [32].

## MATERIALS AND METHODS

The initial overall goal of this work was to assess and compare different approaches to patient-level retrieval by developing a “gold standard” test collection consisting of the three usual components of a Cranfield-style IR collection [15]: records – in this case patient-level medical records, topics – representations of cohorts to be recruited for clinical studies, and relevance judgements – expert determination of which records were relevant to which topics. Our initial plan was to develop the test collection and apply the methods found to work effectively by research groups in the TREC Medical Records Track. However, upon finding the results for numerous topics applied to this data performed sub-optimally, we also developed and evaluated structured Boolean queries, including with additional relevance judgments on a subset of topics.

### Record Collection

As noted in our earlier paper, the patient records originated from OHSU’s Epic (Verona, WI) EHR and were transformed and loaded (without any modification of the underlying structured and textual data) to a research data warehouse [31]. The study protocol to use the records was approved by the OHSU Institutional Review Board (IRB00011159). To be included in the corpus, patients had to have at least three primary care encounters between January 1, 2009 and December 31, 2013, inpatient or outpatient, with at least five text note entries. This was done to ensure that records would more likely be comprehensive of their care as opposed to a patient referred to the academic medical for a single consultation.

Both structured and unstructured data were included in the collection. Document types included demographics, vitals, medications (administered, current, ordered), hospital and ambulatory encounters with associated attributes and diagnoses, clinical notes, problem lists, laboratory and microbiology results, surgery and procedure orders, and result comments. A unique medical record number was used to link the different document types, and each document type could contain multiple data fields. The collection contained a total of 99,965 unique patients and 6,273,137 associated unique encounters. It originated in a relational database but was extracted into XML format for loading into the open-source IR platform Elasticsearch (v1.7.6) for our experiments.

### Topics

The 56 topics used for this research were developed from five sources by OHSU and Mayo Clinic as described in our previous paper [31]. From OHSU, 29 topics were selected from research study data requests submitted by clinical researchers to the Oregon Clinical and Translational Research Institute (OCTRI). From Mayo Clinic, topics were modeled after two patient cohorts found in the Mayo Research Data Warehouse, five patient cohorts in the Phenotype KnowledgeBase (PheKB), nine patient cohorts in the Rochester Epidemiology Project (REP), and 12 patient cohorts based on presence of quality measures from the National Quality Forum (NQF).

Each topic was expressed at three levels of detail, with the complete list in Supplementary Appendix 1:

A. Summary statement – 1-3 sentences
B. Illustrative clinical case
C. Brief summary plus structured inclusion and exclusion criteria for demographics, diagnoses, medications and other attributes

### Initial Runs

As is typically done in Cranfield-style IR experiments, we performed a number of different runs consisting of the text of the topic representation submitted to the ElasticSearch system, which generated ranked output that we limited to 1000 patients per topic. We varied different parameters for different runs by topic representation, text subset, aggregation method, and retrieval model. For the latter, we used a number of common ranking approaches implemented in ElasticSearch and known to be successful both in the TREC Medical Records Track and IR systems generally:

- BM25, also known as Okapi [33]
- Divergence from randomness (DFR) [34]
- Language modeling with Dirichlet smoothing (LMDir) [35]
- Default Lucene scoring, based on the term frequency-inverse document frequency (TF*IDF) model [36]

We performed 48 runs representing all permutations of the following query parameters as described in our previous paper. These representations formed the basis for all queries created for this paper, both manual and automated and include (further referencing in this paper by <u>underlined</u> text):

1. Topic Representation – <u>A</u> (summary statement), <u>B</u> (clinical case), or <u>C</u> (detailed criteria)
2. (Text Subset – only clinical <u>notes</u> or <u>all</u> document types (including structured data reporting as text)
3. Aggregation Method – patient relevance score calculated by summation (<u>sum</u>) of all documents or by maximum (<u>max</u>) value
4. Retrieval Model – <u>BM25</u>, <u>DFR</u>, <u>LMDir</u>, or <u>Lucene</u>

### Relevance Assessment

The relevance assessments were carried out based on the principles discussed in our previous paper [31]. The initial pools for relevance judging were generated in a similar manner to TREC challenge evaluations, where results from different runs were pooled by selecting from all runs for a given topic the top 15 ranked patients and then randomly selecting 25% of the next 85 (21 patients) and 1% of the next 900 (9 patients). The process of relevance judging used the locally developed Patient Relevance Assessment Interface (PRAI) [31]. This system tracked the judgements in a PostgreSQL database and interfaced with the EHR data that was loaded into Elasticsearch. Patient pools for topics were selected for judging and loaded into PRAI, where all document types could be searched by medical experts to determine patient-level relevance for the topic. Document-level sub-relevance could also be assigned in the system. Patients could be assigned one of three levels of relevance: definitely relevant, possibly relevant, or not relevant. For retrieval performance metrics, both definitely and possibly relevant patients were considered relevant, since the use case motivating aimed to identify patients who were likely to be candidates for inclusion in clinical studies, and the number of definite plus possibly relevant patients was typically not vastly larger than would be desired for a clinical trial.

### Assessment of Initial Retrieval Results

We used the standard trec_eval program [37] to generate retrieval results for the 48 runs. Because our queries did not exhaustively assess all possible approaches to retrieval, we opted to use the B-Pref measure for results, based on its common usage for IR evaluation when relevance judging is considered to be incomplete [38]. B-Pref is a measure of how many relevant patients were retrieved, in the ranked lists, ahead of the non-relevant patients in the interval [0,1]. The distribution of B-Pref was evaluated across all 56 topics for each of the 48 runs separately. The intent of this analysis was to assess differences in performance between run parameter settings, and variance within each setting across the 56 topics. We also evaluated the distribution across all of the 48 runs for each of the 56 topics separately to assess differences across topics and variance within each topic across the 48 runs.

As noted in the Results section, the results from these runs were substantially lower than comparable methods applied in the TREC Medical Records Track. This led us to perform additional methods described in the rest of this section that included use of structured Boolean queries on a subset of topics.

### Structured Queries

Because the word-based query methods that had worked well for the TREC Medical Records Track performed less successfully with this data, we constructed structured Boolean queries for the 56 topics in an iterative manner by one of the authors with clinical experience (SRC). These queries were based on Topic Representation C, which contained structured data and some free text. These queries were allowed to search all document types that we loaded into Elasticsearch. Since these were structured queries, we did not rank patients returned, and all patients returned were kept in the final query results. Patients retrieved could have been part of the word-based retrieval pools and thus known to be relevant or not, or have not been judged. In addition, some patients not retrieved by the structured queries could have been relevant from retrieval and judgments in the word-based pools. Standard to their definitions, recall for each query was measured as patients retrieved and relevant / patients known to be relevant and precision was measured as patients retrieved and relevant / patients retrieved. We also measured patients retrieved who had not been judged for relevance in the initial pool.

### Additional Relevance Assessment for Ten Selected Topics

As we discovered that a number of patients retrieved by the structured queries had not been retrieved by the word-based queries and therefore not judged, we selected ten topics for additional relevance judging of patients returned by the structured queries. These included topics 2, 7, 9, 17, 32, 33, 42, 44, 48, and 52. To build on previous work done in our group, we used five topics that had been selected randomly for this previous research [39], while the second five topics were selected for diversity in all five of our sources for topic definitions (OHSU, Mayo, PheKP, REP, NQF). The second five were also selected based on a higher likelihood to be seen in clinical practice (based on clinician judgement), as compared to other topics in the list of 56.

For these ten topics our intent was to judge the entire list of patients retrieved by the structured queries. To compare the structured queries to the word-based queries we used simple precision and recall. B-Pref was not an appropriate measure since the judged structured query patient pools were not ranked. For recall, we combined the relevant patients found in both the structured judged pools and the word-based judged pools. We counted patients judged as definitely or possibly relevant as relevant for all analyses. We also measured relevant patients retrieved in the word-based queries but not in the structured queries.

## RESULTS

### Word-based Query Results

Per the Cranfield approach, we performed standard batch runs for the 48 permutations of topic representation, subset, aggregation method, and retrieval model. For relevance judging, the results were pooled by topic. Relevance assessing of patients was done by a physician who took around 30-40 hours per topic. Table 1 shows the number, source, summary, and distribution of relevance judgments for a sampling of 10 topics, with the full table of all 56 topics in Supplementary Appendix 2. One topic had no definite or possibly relevant patients and was excluded from further analysis (25). We used the trec_eval program to include each topic for each run, along with the relevance judgments, to generate retrieval results for each run.

**Table 1.** A sample of the 56 topics with number, source, summary, and pool size, as described in the text. Also shown are number and percentage for definitely relevant, possibly relevant, and not relevant from the initial relevance assessment process.

| Num | Source | Summary | Pool | Def<br>Rel | % | Poss<br>Rel | % | Not<br>Rel | % |
| --- | --- | --- | --- | --- | --- | --- | --- | --- | --- |
| 2 | OHSU | Adults with IBD who haven't had GI surgery | 684 | 63 | 9.2% | 4 | 0.6% | 617 | 90.2% |
| 7 | OHSU | Hereditary hemorrhagic telangiectasia | 695 | 15 | 2.2% | 0 | 0.0% | 680 | 97.8% |
| 9 | OHSU | Children with focal epilepsy with partial seizures | 687 | 31 | 4.5% | 13 | 1.9% | 643 | 93.6% |
| 17 | OHSU | RA on MTX w/o biologic DMARD | 704 | 20 | 2.8% | 0 | 0.0% | 684 | 97.2% |
| 32 | PheKB | ACE inhibitor-induced cough | 700 | 40 | 5.7% | 0 | 0.0% | 660 | 94.3% |
| 33 | PheKB | Children with ADHD on CNS stimulant | 732 | 112 | 15.3% | 0 | 0.0% | 620 | 84.7% |
| 42 | NQF | Elderly patients with dementia on antipsychotic medication | 731 | 24 | 3.3% | 0 | 0.0% | 707 | 96.7% |
| 44 | NQF | COPD with potentially avoidable complication | 680 | 38 | 5.6% | 0 | 0.0% | 642 | 94.4% |
| 48 | REP | Stroke after first MI | 698 | 5 | 0.7% | 0 | 0.0% | 693 | 99.3% |
| 52 | REP | Cataract surgery and prior SSRI use | 737 | 23 | 3.1% | 13 | 1.8% | 701 | 95.1% |

The highest overall performing run was b.notes.max.LMDir, with a mean B-Pref of 0.167. Very close to this run were two variations of the Retrieval Model: b.notes.max.DFR, and b.notes.max.Lucene, although b.notes.max.BM25 scored lower. At the other end of performance, the a.notes.sum.BM25 run had a mean B-Pref of 0.106. Figure 1 depicts the median and distribution of B-Pref for all 48 runs across all 56 topics (Figure 1).

**Figure 1.**
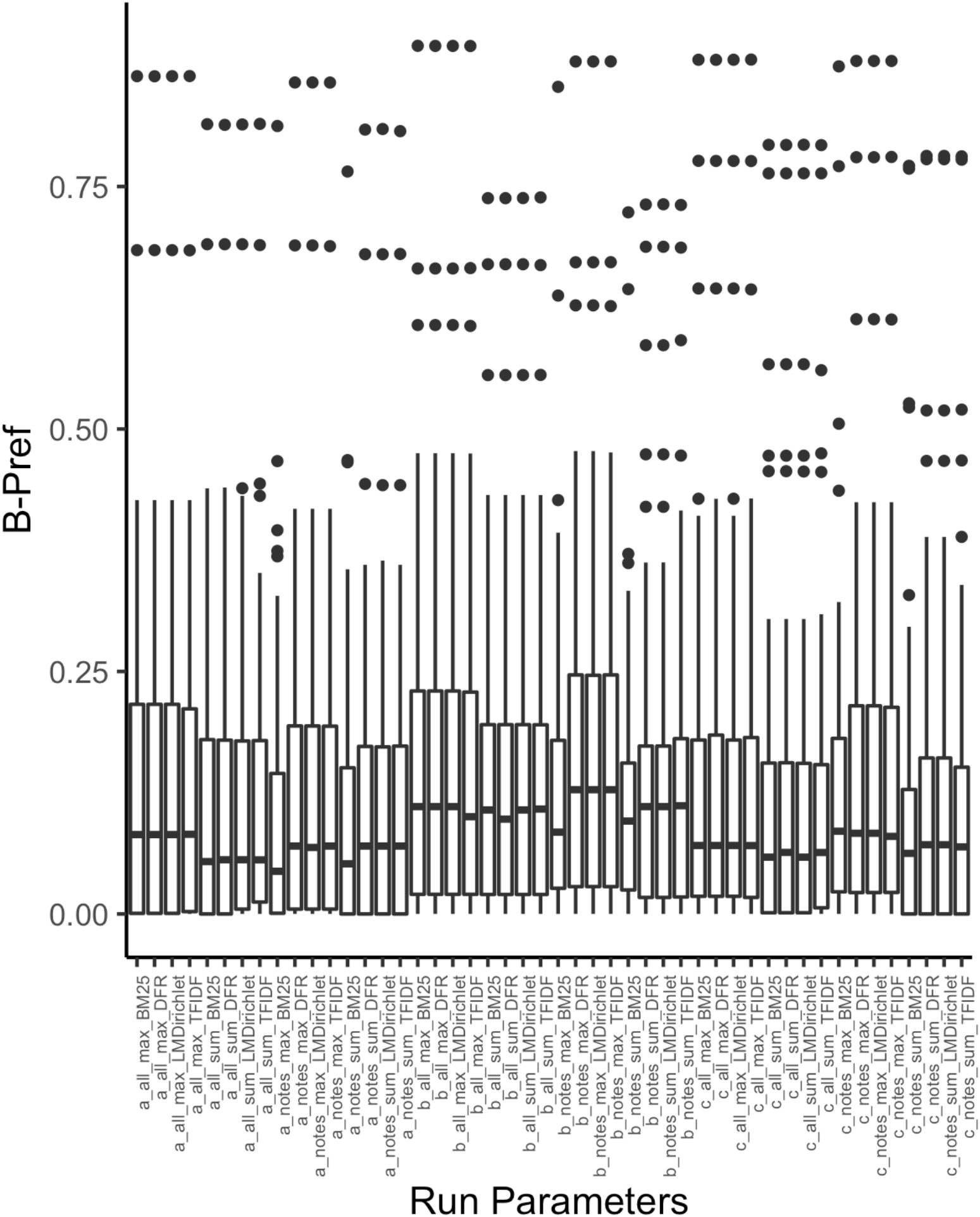
B-Pref distributions for topics within each run. Box ends represent the upper and lower quartile values and whiskers extend 1.5 times the interquartile range. Data points beyond the end of the whiskers are values for individual topics outside the whiskers. The parameter settings are ordered hierarchically first by Topic Representation (A, B, C), then Text Subset (all, notes), then Aggregation Method (max, sum) and finally the Retrieval Model (BM25, DFR, LMDirichlet, TFIDF).

There were several performance grouping patterns seen within the four parameters. Overall, Topic Representation B performed better than the other two representations. This representation was only comprised of text, but included a detailed individual case description, along with summary description. There was a tendency for the Retrieval Model BM25 to perform poorer than the other models, primarily with the Text Subset notes, which was comprised of a more limited use of available document types. There was a trend for the Aggregation Method sum to have lower performance than the method max.

As is commonly seen in IR experiments, the distribution of topics was spread widely (Figure 2). The highest mean B-Pref was for topic 50 (0.895), while 11 topics had essentially a mean B-Pref of 0.0 (i.e., most runs retrieved no relevant patients). Two topics consistently had the top two highest values for B-Pref for all parameter combinations within Topic Representation A and C, topics 50 and 28. For Topic Representation B, topic 50 was also consistently in the top two extreme B-Pref values along with topic 47. These topics did not have complex temporal conditions, medication requirements or surgery inclusions or exclusions, and only required relatively straightforward inclusion/exclusion lists of medical conditions, lab and radiology tests, and demographics.

**Figure 2.**
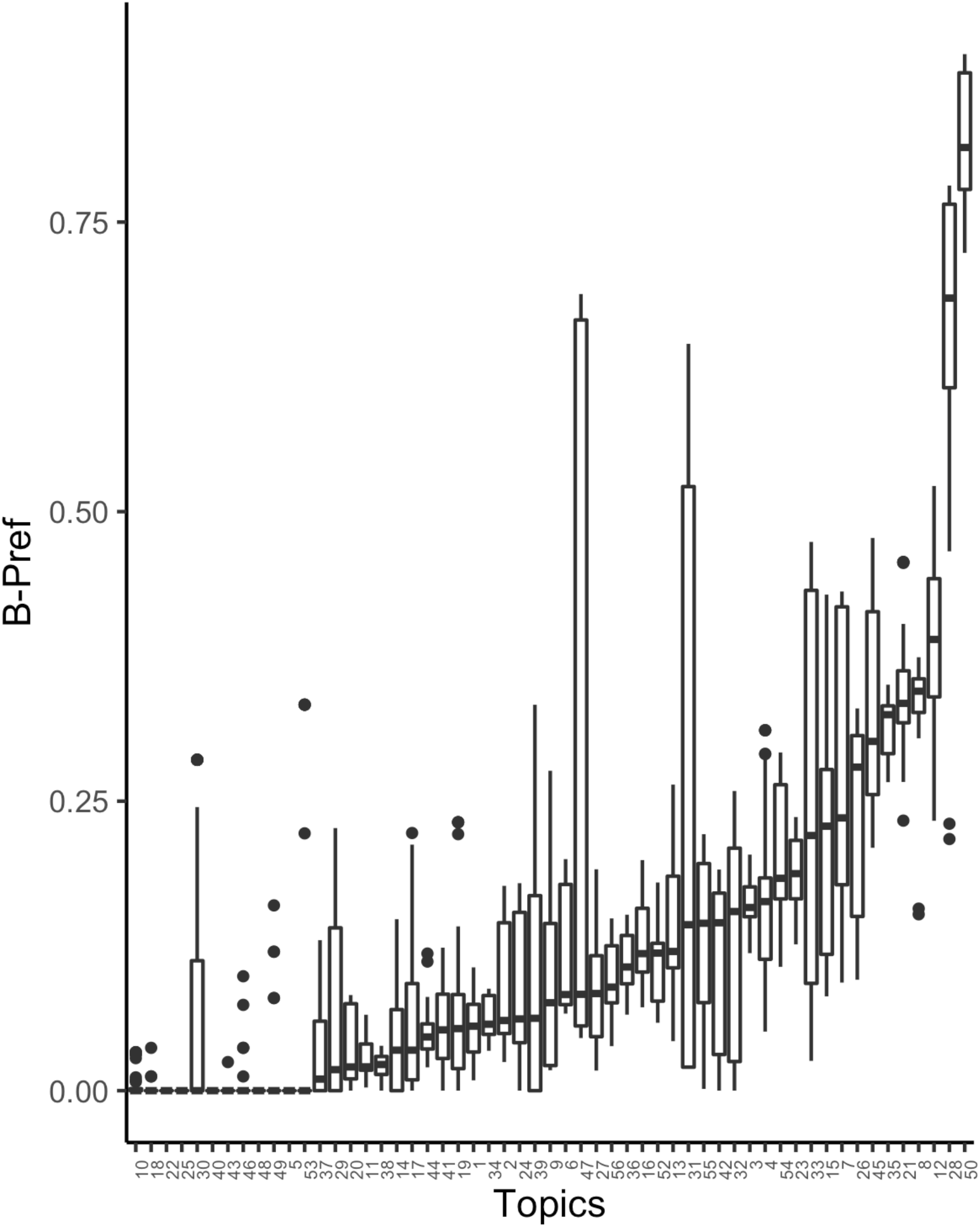
B-Pref distributions for parameter combinations within each topic. Box ends represent the upper and lower quartile values and whiskers extend 1.5 times the interquartile range. Data points beyond the end of the whiskers are values for parameter combinations outside the whiskers. Boxplots are ordered by median B-Pref values.

B-Pref distributions of the 48 parameter combinations (runs) within each topic varied widely in range and shape. Topics 31 and 47 were distinctive, showing much greater variation in performance across parameter settings than the other topics. This variation was entirely due to large differences between Topic Representations. There was very little performance variation for these two topics across the other parameter combinations within each representation.

### Structured Queries

For each topic, we calculated simple recall and precision on the output of each structured Boolean query (Boolean queries are typically not ranked) using the relevance judgments for the word-based query pools. As with the word-based queries, a patient was considered relevant if rated definitely or possibly relevant. Table 2 shows an example structured Boolean query for Topic 7. Recall for the structured queries varied widely across topics (Figure 3). There was 100% recall of word-based query relevant patients on 8 of the 56 topics, greater than 50% recall on 35 of the 56 topics, less then 50% recall on 13 of the 56, one topic (48) with no recall of relevant patients, and two topics with no retrieval at all (22, 25).

**Table 2.**
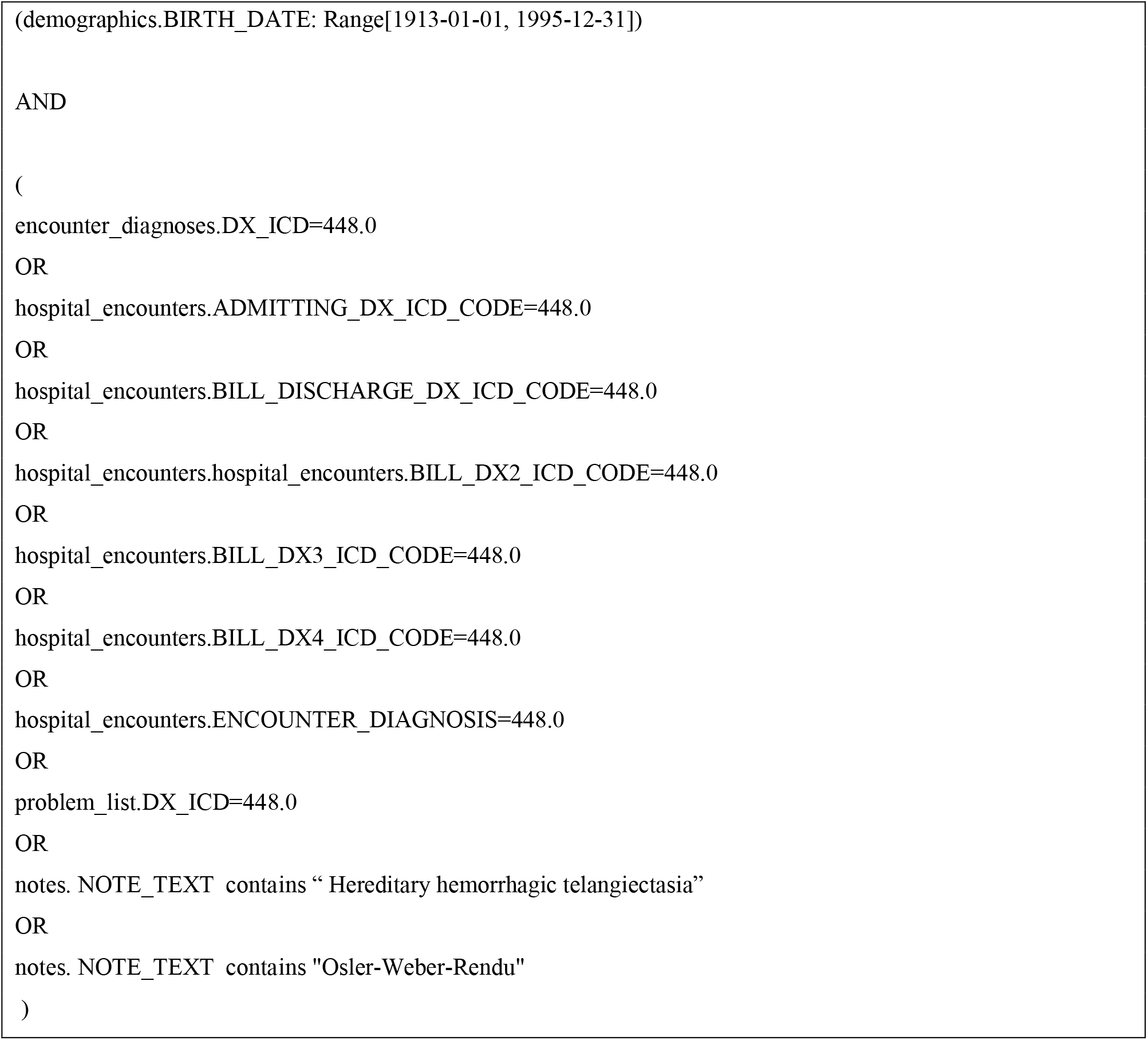
Structured Boolean query for Topic 7: Adults 18-100 years old who have a diagnosis of hereditary hemorrhagic telangiectasia (HHT), which is also called Osler-Weber-Rendu syndrome.

**Figure 3.**
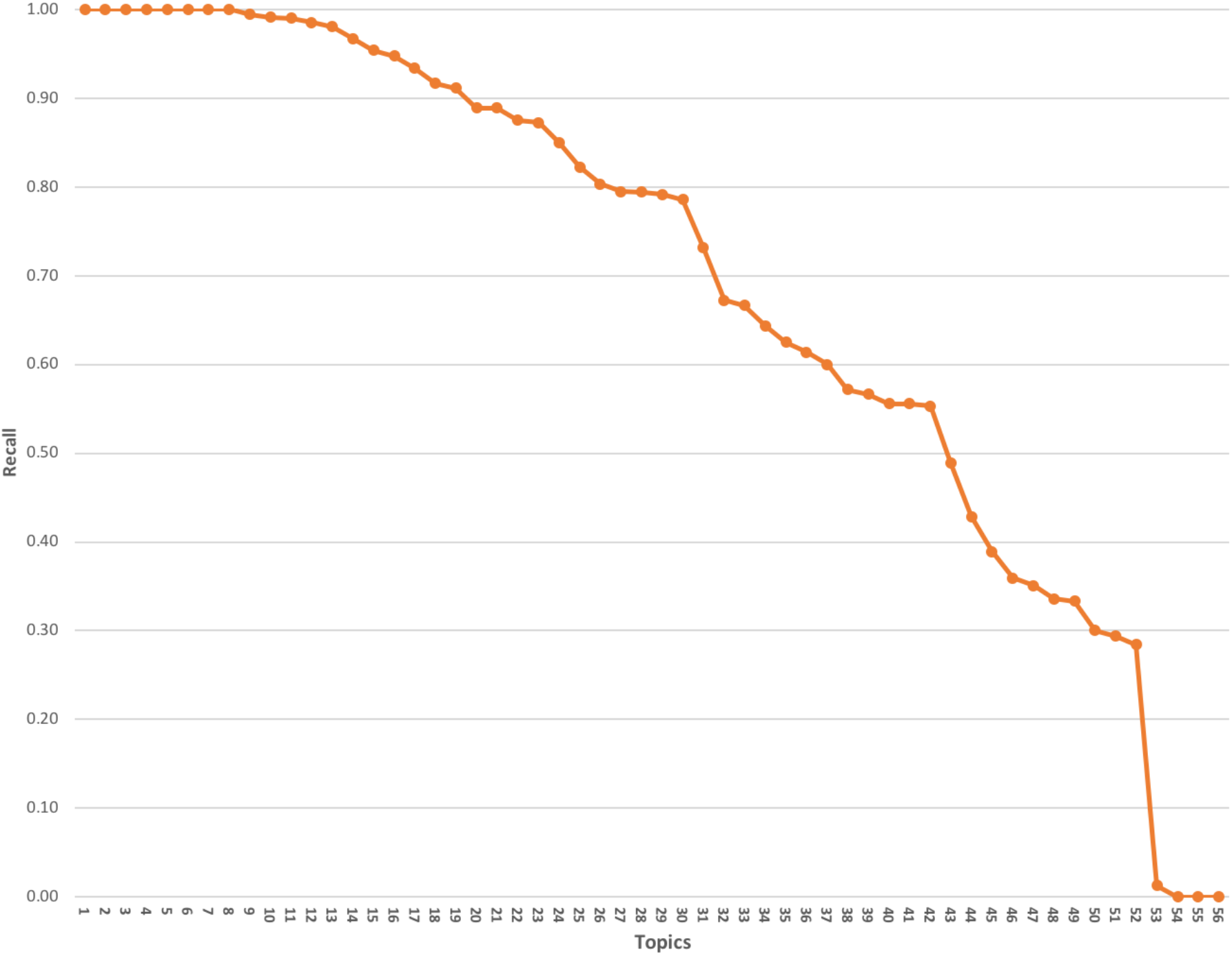
Recall of relevant patients from word-based query pools by structured queries, ordered by recall for each topic.

Precision likewise varied widely across topics for the structured queries, (Figure 4). The structured queries outperformed the word-based queries in precision for all topics except 48. Again, topics 22 and 25 did not return any relevant patients. Three topics had 100% precision (29, 34, 46).

**Figure 4.**
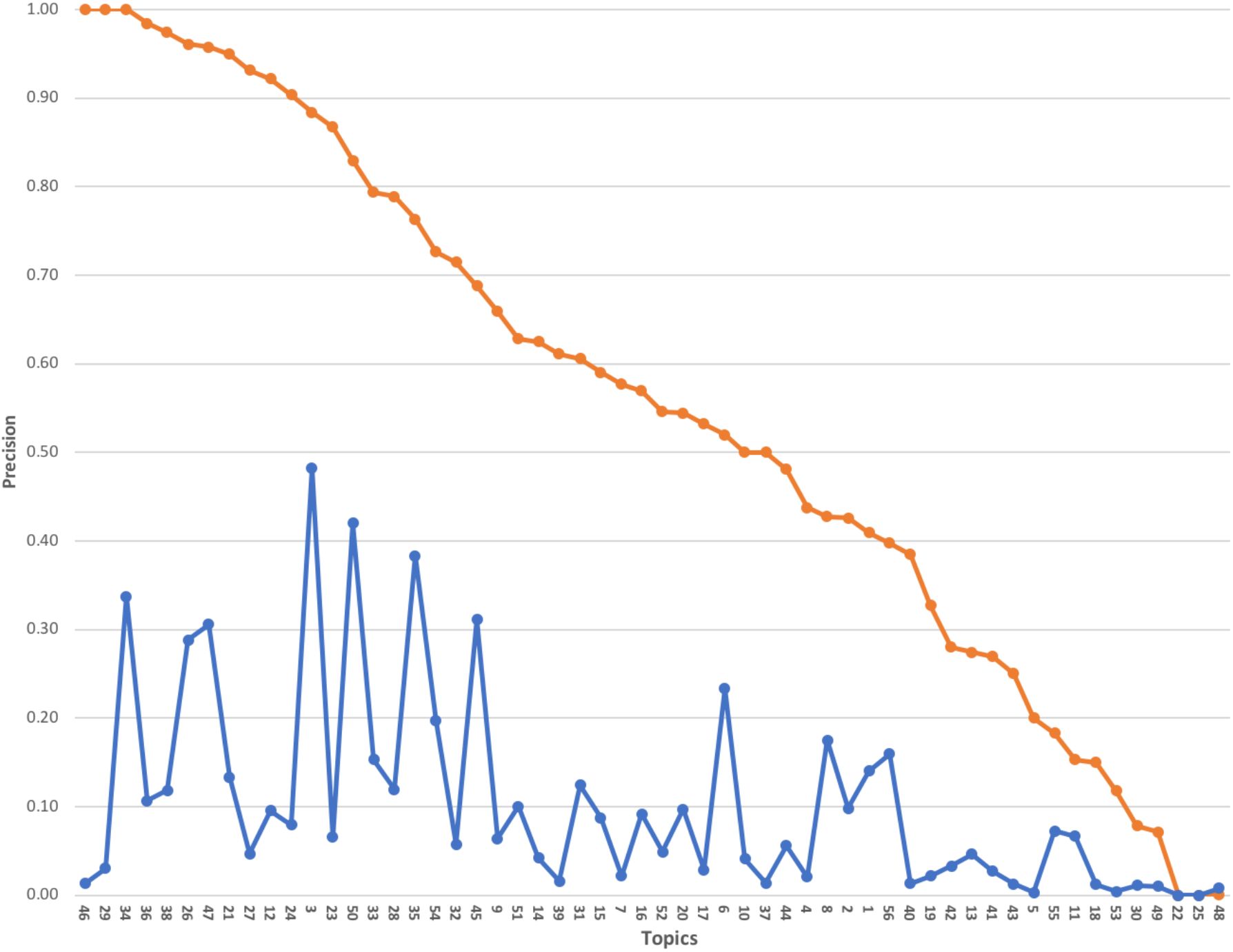
Precision for structured queries (red line) and word-based judged pools (blue line), ordered by structured query precision for each topic.

### Topics with Expanded Relevance Judgements for the Structured Queries

Because the structured queries retrieved patients who had not been retrieved by the word-based queries, we did additional relevance judging for ten selected topics. Due to the large number of patients returned for topic 2 by the structured query (2,578), only a random sample of 750 patients was judged.

Although the structured queries had higher recall than the word-based queries for all ten topics, these queries did not achieve complete recall of all of the relevant patients for nine of the ten topics (Figure 5). The numbers of relevant patients found only in word-based queries was relatively low, compared to the total number of relevant patients (Table 3). This explains the larger number of missed relevant patients for this topic. The structured queries had higher precision for all ten topics (Figure 6). For topic 52, all patients retrieved by the structured query were judged relevant.

**Table 3.** Ten topics with additional relevance judgments for results from structured Boolean queries. The structured queries retrieved additional patients who were judged for relevance, allowing calculation of recall and precision for these queries as well as determination of numbers found by the word-based queries but missed by the structured queries.

| Topic | Structured query | Word-based | Structured query | Structured query relevant | Recall for structured query | Precision for | Structured query |
| --- | --- | --- | --- | --- | --- | --- | --- |

|  | patients<br>retrieved | query<br>relevant | added<br>relevant | and<br>retrieved |  | structured<br>query | relevant<br>missed |
| --- | --- | --- | --- | --- | --- | --- | --- |
| 2 | 750 | 67 | 490 | 438 | 0.89 | 0.58 | 52 |
| 7 | 50 | 15 | 24 | 24 | 1.00 | 0.48 | 0 |
| 9 | 357 | 44 | 190 | 173 | 0.91 | 0.48 | 17 |
| 17 | 110 | 20 | 112 | 109 | 0.97 | 0.99 | 3 |
| 32 | 390 | 40 | 368 | 353 | 0.96 | 0.91 | 15 |
| 33 | 1092 | 112 | 983 | 982 | 1.00 | 0.90 | 1 |
| 42 | 347 | 24 | 347 | 344 | 0.99 | 0.99 | 3 |
| 44 | 378 | 38 | 266 | 264 | 0.99 | 0.70 | 2 |
| 48 | 68 | 5 | 37 | 32 | 0.86 | 0.47 | 5 |
| 52 | 133 | 36 | 157 | 133 | 0.85 | 1.00 | 12 |

**Figure 5.**
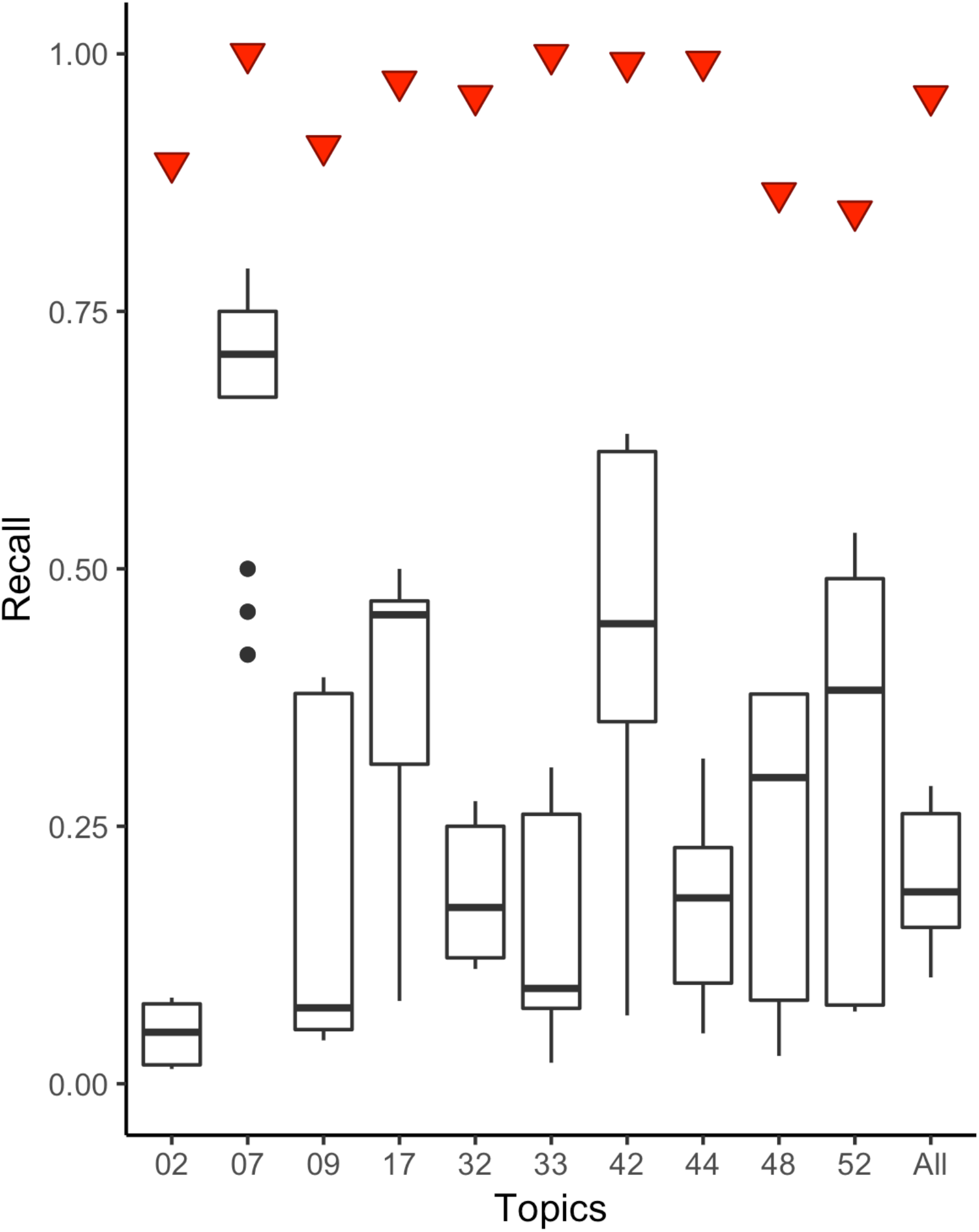
Recall distributions for ten selected topics based on combined full structured-query relevance judged pools. Red triangles are the values for the structured queries while the box and whisker plots contain the distributions for word-based queries with the original 48 different parameters.

**Figure 6.**
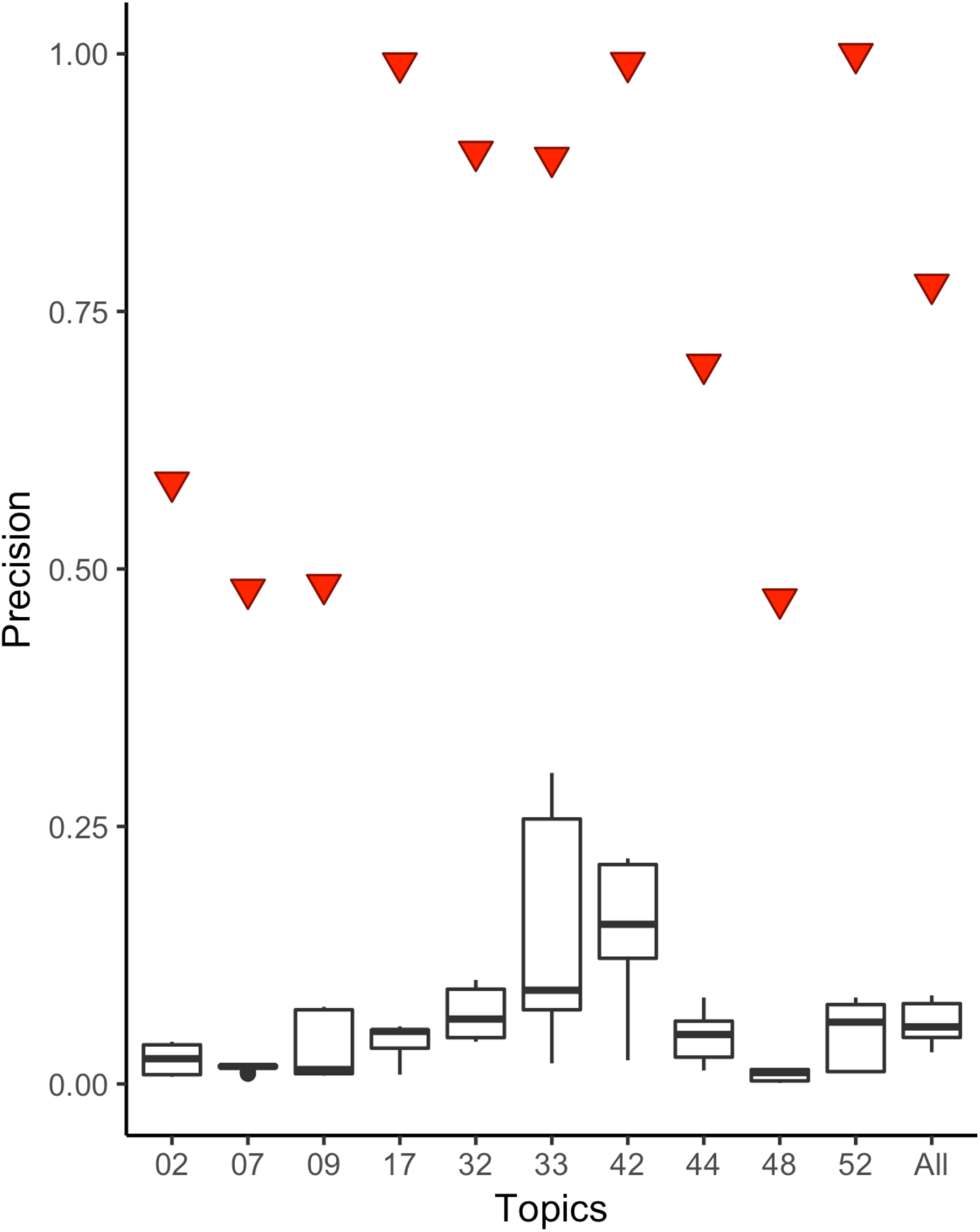
Precision distributions for ten selected topics based on combined full structured-query relevance judged pools. Red triangles are the values for the structured queries while the box and whisker plots contain the distributions for word-based queries with the original 48 different parameters.

## DISCUSSION

We set out to begin this work using word-based query methods that performed well for the TREC Medical Records Track. Our results did not achieve the performance we expected (Figure 1). Overall, the best results were achieved with the Topic Representation of the illustrative clinical case formulation (B), with small further improvements for using Text Subset all and Aggregation Method max. Within our results, we observed variation common to IR challenge evaluations. Although the overall differences were modest, there was consistently higher values for Topic Representation B. Likewise, there was small benefit for Aggregation Method sum vs. max. For combinations of parameters, the Retrieval Model BM25 performed worse than the other three. To the extent that these results are generalizable, clinical case formulations are the best query type among word-based methods for the patient cohort discovery task.

Also common to IR challenge evaluation results, reflecting the adage that means and medians can obscure variations, there was a large difference in retrieval performance by topic. As seen in Figure 2, about ten had very poor performance while two had very high performance across all retrieval methods. There was also a substantial range of performance within a number of individual topics.

In the effort to improve our results, we reformulated our queries using structured Boolean approaches developed iteratively. Because pure Boolean queries do not rank their output, we could not directly compare our results with the word-based queries. Instead, we measured standard recall and precision based on the relevance judgments made for patients retrieved by the word-based methods. The results for the structured queries were much better, with a median recall of 0.86 and eight topics having recall of 1.0 (Figure 3). There were likewise 13 topics with recall under 0.4 and a couple near zero. Precision was not associated with recall for the topics but did vary almost linearly from 1.0 to 0.0 across the topics (Figure 4).

One concern for the structured queries was the use of the relevance judgments only from the word-based query results. As such, we performed additional relevance judgments based on the structured query retrieval for 10 topics. This not only would give us a more realistic picture of the performance of these topics, but also identify additional patients for relevance judgment for the word-based queries. After the additional judgments, we found that the structured queries had much higher recall than the word-based queries (Figure 5) as well as much higher precision (Figure 6), which was also been found in comparable experiments from Mayo Clinic [39]. As precision is sometimes conceptualized as “number needed to read” [40], the higher precision for the structured queries means fewer patients would need to be assessed to identify candidates for clinical studies.

There were a number of limitations to this work. Our records were limited to a single academic medical center. There are many additional retrieval methods we could have assessed, but we would not have the resources to carry out the additional relevance judgments required as those additional methods would add new patients to be judged. Finally, there is a global limitation to work with EHR data for these sorts of use cases in that raw, identifiable patient data is not easily sharable such that other researchers could compare their systems and algorithms with ours using our data, although they could apply our methods to their own data.

## CONCLUSIONS

Cohort retrieval is commonly offered by many medical centers with EHR systems but is poorly understood and, with current approaches, still labor-intensive. Automated methods can likely improve performance of systems and reduce time taken to identify definitely relevant patients, although manual crafting of Boolean queries showed much better performance in this research. Challenges to developing and evaluating IR methods for this use case include the resources required to perform relevance judgments and the nature of such highly private data that makes their comparison across different research groups difficult. Our future work will continue to develop methods that show promise and evaluate them with real-world topics and relevance judgments in our data. We also plan to classify topic characteristics and assess their role in retrieval performance.

## Data Availability

The data used for this study is protected health information that came from the electronic health record system at Oregon Health & Science University, so cannot be made publicly available.

## ACKNOWLEDGMENTS

This work was supported by NIH Grant 1R01LM011934.

## REFERENCES

1. Murphy, S., et al., Current state of information technologies for the clinical research enterprise across academic medical centers. Clinical and Translational Science, 2012. 5: p. 281–284.

2. Obeid, J., et al., A survey of practices for the use of electronic health records to support research recruitment. Journal of Clinical and Translational Science, 2017. 1: p. 246–252.

3. Sholle, E., et al., A scalable method for supporting multiple patient cohort discovery projects using i2b2. Journal of Biomedical Informatics, 2018. 84: p. 179–183.

4. Visweswaran, S., et al., Accrual to Clinical Trials (ACT): a clinical and translational science award consortium network. JAMIA Open, 2018. 1(2): p. 147–152.

5. Topaloglu, U. and M. Palchuk, Using a federated network of real-world data to optimize clinical trials operations. JCO Clinical Cancer Informatics, 2018. 2: p. 1–10.

6. Ni, Y., et al., Increasing the efficiency of trial-patient matching: automated clinical trial eligibility pre-screening for pediatric oncology patients. BMC Medical Informatics & Decision Making, 2015. 15: p. 28.

7. Ni, Y., et al., Automated clinical trial eligibility prescreening: increasing the efficiency of patient identification for clinical trials in the emergency department. Journal of the American Medical Informatics Association, 2015. 22: p. 166–178.

8. Ni, Y., et al., A real-time automated patient screening system for clinical trials eligibility in an emergency department: design and evaluation. JMIR Medical Informatics, 2019. 7(3): p. e14185.

9. Chapman, W., et al., Overcoming barriers to NLP for clinical text: the role of shared tasks and the need for additional creative solutions. Journal of the American Medical Informatics Association, 2011. 18: p. 540–543.

10. Friedman, C., T. Rindflesch, and M. Corn, Natural language processing: state of the art and prospects for significant progress, a workshop sponsored by the National Library of Medicine. Journal of Biomedical Informatics, 2013. 46: p. 765–773.

11. Chapman, W., et al. Creation of a repository of automatically de-identied clinical reports: processes, people, and permission. in Proceedings of the American Medical Informatics Association Clinical Reserach Informatics Summit. 2011. San Francisco, CA.

12. Johnson, A., et al., MIMIC-III, a freely accessible critical care database. Scientific Data, 2016. 3: p. 160035.

13. Voorhees, E. and R. Tong. Overview of the TREC 2011 Medical Records Track. in The Twentieth Text REtrieval Conference Proceedings (TREC 2011). 2011. Gaithersburg, MD: National Institute of Standards and Technology.

14. Voorhees, E. and W. Hersh. Overview of the TREC 2012 Medical Records Track. in The Twenty-First Text REtrieval Conference Proceedings (TREC 2012). 2012. Gaithersburg, MD: National Institute of Standards and Technology.

15. Cleverdon, C. and E. Keen, Factors determining the performance of indexing systems (Vol. 1: Design, Vol. 2: Results). 1966, Aslib Cranfield Research Project: Cranfield, England.

16. Voorhees, E. The TREC Medical Records Track. in Proceedings of the International Conference on Bioinformatics, Computational Biology and Biomedical Informatics. 2013. Washington, DC.

17. Zhu, D., et al., Using large clinical corpora for query expansion in text-based cohort identification. Journal of Biomedical Informatics, 2014. 49: p. 275–281.

18. Goodwin, T. and S. Harabagiu, Learning relevance models for patient cohort retrieval. JAMIA Open, 2018. 1: p. 265–274.

19. Sarmiento, R. and F. Dernoncourt, Improving Patient Cohort Identification Using Natural Language Processing, in Secondary Analysis of Electronic Health Records, Anonymous, Editor. 2016, Springer: Cham, Switzerland. p. 405–417.

20. Glicksberg, B., et al. Automated disease cohort selection using word embeddings from electronic health records. in Pacific Symposium on Biocomputing. 2018.

21. Stubbs, A., et al., Cohort selection for clinical trials: n2c2 2018 shared task track 1. Journal of the American Medical Informatics Association, 2019. 26: p. 1163–1171.

22. Ateya, M., B. Delaney, and S. Speedie, The value of structured data elements from electronic health records for identifying subjects for primary care clinical trials. BMC Medical Informatics & Decision Making, 2016. 16: p. 1.

23. Kang, T., et al., EliIE: an open-source information extraction system for clinical trial eligibility criteria. Journal of the American Medical Informatics Association, 2017. 24: p. 1062–1071.

24. Zhang, K. and D. Demner-Fushman, Automated classification of eligibility criteria in clinical trials to facilitate patient-trial matching for specific patient populations. Journal of the American Medical Informatics Association, 2017. 24: p. 781–787.

25. Yuan, C., et al., Criteria2Query: a natural language interface to clinical databases for cohort definition. Journal of the American Medical Informatics Association, 2019. 26: p. 294–305.

26. Wu, H., et al., SemEHR: a general-purpose semantic search system to surface semantic data from clinical notes for tailored care, trial recruitment, and clinical research. Journal of the American Medical Informatics Association, 2018. 25: p. 530–537.

27. Gligorijevic, J., et al., Optimizing clinical trials recruitment via deep learning. Journal of the American Medical Informatics Association, 2019: p. Epub ahead of print.

28. Denny, J., L. Bastarache, and D. Roden, Phenome-wide association studies as a tool to advance precision medicine. Annual Review of Genomics and Human Genetics, 2016. 17: p. 353–373.

29. Richesson, R., et al., Clinical phenotyping in selected national networks: demonstrating the need for high-throughput, portable, and computational methods. Artificial Intelligence in Medicine, 2016. 71: p. 57–61.

30. Robinson, J., et al., Defining phenotypes from clinical data to drive genomic research. Annual Review of Biomedical Data Science, 2018. 1: p. 69–92.

31. Wu, S., et al., Intra-institutional EHR collections for patient-level information retrieval. Journal of the American Society for Information Science & Technology, 2017. 68: p. 2636–2648.

32. Wang, Y., et al., Test collections for electronic health record-based clinical information retrieval. JAMIA Open, 2019. 2: p. 360–368.

33. Robertson, S. and S. Walker. Some simple effective approximations to the 2-Poisson model for probabilistic weighted retrieval. in Proceedings of the 17th Annual International ACM SIGIR Conference on Research and Development in Information Retrieval. 1994. Dublin, Ireland: Springer-Verlag.

34. Amati, G. and C. VanRijsbergen, Probabilistic models of information retrieval based on measuring the divergence from randomness. ACM Transactions on Information Systems, 2002. 20: p. 357–389.

35. Zhai, C. and J. Lafferty, A study of smoothing methods for language models applied to information retrieval. ACM Transactions on Information Systems, 2004. 22: p. 179–214.

36. Salton, G. and C. Buckley, Term-weighting approaches in automatic text retrieval. Information Processing and Management, 1988. 24: p. 513–523.

37. Harman, D., Information Retrieval Evaluation. 2011, San Rafael, CA: Morgan & Claypool.

38. Buckley, C. and E. Voorhees. Retrieval evaluation with incomplete information. in Proceedings of the 27th Annual International ACM SIGIR Conference on Research and Development in Information Retrieval. 2004. Sheffield, England: ACM Press.

39. Liu, S., et al., CREATE: cohort retrieval enhanced by analysis of text from electronic health records using OMOP common data model. arXiv.org, 2019.

40. Bachmann, L., et al., Identifying diagnostic studies in MEDLINE: reducing the number needed to read. Journal of the American Medical Informatics Association, 2002. 9: p. 653–658.

